# Dysbiosis in pre-cancerous mucosa may drive immune dysfunction – a study of immune system: microbiota interaction in familial adenomatous polyposis

**DOI:** 10.1101/2021.05.05.21256687

**Authors:** Alistair Noble, Lydia Durant, Stella M Dilke, Ripple Man, Isabel Martin, Roshani Patel, Lesley Hoyles, Edward T Pring, Andrew Latchford, Susan K Clark, Simon R Carding, Stella C Knight

**Affiliations:** Gut Microbes & Health Programme, Quadram Institute Bioscience, Norwich, UK; Antigen Presentation Research Group, Imperial College London, Northwick Park & St. Mark’s Campus, Harrow, UK; The Polyposis Registry, St. Mark’s Hospital, London North West University Healthcare NHS Trust, Harrow, UK; Department of Surgery and Cancer, Imperial College London, UK; Department of Biosciences, Nottingham Trent University, Nottingham, UK; Norwich Medical School, University of East Anglia, Norwich, UK

**Keywords:** familial adenomatous polyposis, colorectal cancer, resident memory T-cells, intraepithelial lymphocytes, intestinal microbiota

## Abstract

**Objectives:** Familial adenomatous polyposis (FAP) is a condition caused by a constitutional pathogenic variant of the adenomatous polyposis coli (*APC*) gene that results in intestinal adenoma formation and colorectal cancer (CRC), necessitating pre-emptive colectomy. We sought to examine interaction between the mucosal immune system and commensal bacteria in FAP to test for immune dysfunction that might accelerate tumorigenesis.

**Methods:** Colonic biopsies were obtained from macroscopically normal mucosal tissue from 14 healthy donors and 13 patients with FAP during endoscopy or from surgical specimens. Intraepithelial and lamina propria lymphocytes were phenotyped. Intraepithelial microbes were labelled with anti-IgA/IgG and analyzed by flow cytometry.

**Results:** Proportions of resident memory CD103-expressing CD8^+^ and γδ T cell receptor^+^ intraepithelial lymphocytes were dramatically reduced in both left and right colon of patients with FAP compared to healthy controls. In lamina propria, T-cells expressed less CD103 and CD4^+^ CD103^+^ cells expressed less CD73 ectonucleotidase. IgA coating of epithelia-associated bacteria, IgA^+^ peripheral B cells and CD4 T-cell memory responses to commensal bacteria were increased in FAP.

**Conclusions:** Loss of resident memory T-cells and γδ T-cells in mucosal tissue of patients with FAP accompanies intestinal microbial dysbiosis previously reported in this pre-cancerous state and suggests impaired cellular immunity and tumor surveillance. This may lead to barrier dysfunction, possible loss of regulatory T-cell function and excess IgA antibody secretion. Our data are the first to implicate mucosal immune dysfunction as a contributing factor in this genetically driven disease and identify potentially critical pathways in the etiology of CRC.

## Introduction

The intestinal microbiota is altered (dysbiosis) in patients with colorectal cancer (CRC)^1, 2^, but how this might contribute to tumor formation, via interactions with the epithelium and the large numbers of lymphocytes resident in mucosal tissue, is unknown. CRC is accompanied by parainflammation^3^ which may be driven by enhanced exposure of the immune system to microbiota constituents. Our laboratory has recently demonstrated a loss of resident memory T cells (Trm) and γδ T cells in the extra-tumoral tissue of patients with CRC, accompanied by altered immunity to commensal intestinal bacteria and a B cell activation signature in peripheral blood^4^. Whether these factors have a direct causative role in tumor initiation is unclear, but this could be elucidated through studies of pre-cancerous states.

Familial adenomatous polyposis (FAP) is a condition caused by a constitutional pathogenic variant in *APC*, which predisposes individuals to intestinal adenoma formation and CRC^5^. It affects around 1 in 7000 people^6^, who develop up to hundreds or thousands of adenomas in the large bowel, leading almost invariably to CRC by their fourth to fifth decade unless prophylactic surgery is performed^7^. The location of the constitutional *APC* pathogenic variant can predict the colorectal phenotype with a pathogenic variant in the mutation cluster region (codon 1250 to 1450) predicting higher adenoma number compared to those at the 3’ or 5’ of the *APC* gene. *APC* acts as a tumor suppressor gene and is involved in Wnt signaling, cytoskeleton organisation, and T cell immune synapse formation^8,9^, which controls T cell cytokine production and development of regulatory T cells^10^. Development of intestinal tumors in *APC* mutant mice is accompanied by inflammation^11^ and impaired production of the regulatory cytokine IL-10 in intestinal lamina propria T cells^10^.

CRC is associated with intestinal microbial dysbiosis^1^, thought to drive chronic low level or para-inflammation. Maintenance of the barrier between intestinal microbiota and the systemic immune system involves tissue-resident lymphocytes including CD4^+^ and CD8^+^ Trm and γδ T cells^12^. The presence of high numbers of Trm in a variety of tumor tissues is associated with a good prognosis^13^, and induction of Trm by mucosal vaccines is more successful at tumor control in animal models^14^. Thus, Trm appear to suppress both tumor development and inflammation while γδ T cells possess unique function in early tumor surveillance. Intestinal barrier function in patients with FAP is reported to be compromised, with biofilms including *Escherichia coli* and *Bacteroides fragilis* penetrating the mucous barrier^15^. Therefore, by studying immune system: microbiota interaction in human pre-cancerous tissue, we sought to shed light on the factors that increase susceptibility to CRC and for the first time define an immune signature in FAP.

## Materials and Methods

### Study Design

Donors (age 16-80 years) were recruited from St. Mark’s Hospital Polyposis Registry and included those with a diagnosis of FAP (with a confirmed *APC* pathogenic variant) without previous surgery, undergoing colonic surveillance or surgery to remove the large intestine. “Healthy” donors were undergoing investigative endoscopy due to a change in bowel habit or family history of CRC, with no abnormalities observed. Patients were recruited between March 2018 and October 2019; no data were excluded at the end of the study. Clinical and demographic patient characteristics are shown in Table 1. Surgical tissue from the left and right colon and/or 20ml peripheral blood was obtained from all participants. Ethical approval was obtained from the Health Research Authority UK and London-Harrow Research Ethics Committee (study ref 17/LO/1636). Written informed consent was received from participants prior to inclusion in the study.

**Table 1.** Demographic data on study participants.

| Group | HEALTHY | FAP |
| --- | --- | --- |
| Tissue/blood donors: |  |  |
| n | 14 | 13 |
| Male/female | 11/3 | 4/9 |
| Mean age (95% CI) at sampling | 51.3 (43.2-59.3) | 25.7 (18.3-33.1) |
| Biopsy procedure: |  |  |
| Endoscopy |  | 7 |
| Surgery tissue |  | 6 |

### Colonic intraepithelial lymphocytes (IEL), lamina propria lymphocytes (LPL) and intraepithelial microbe (IEM) isolation

For most tissue donors, five left colon and five right colon biopsies were obtained from mucosa with macroscopically normal appearance during routine colonoscopy. Control colonic biopsies (five left colon, five right colon), were obtained from healthy individuals undergoing investigative endoscopy. For six patients with FAP, five left and five right colon mucosal biopsies (10 mg tissue each) were obtained from macroscopically normal (non-adenomatous) appearing mucosa from fresh surgical specimens obtained immediately after surgery. Surgical tissue biopsies were taken by forceps in a manner as comparable as possible to biopsy at endoscopy. Patients undergoing surgery were treated with antibiotics and mechanical bowel preparation (laxatives), as part of routine bowel preparation within the 24-hour period prior to surgery. Individuals undergoing endoscopy received mechanical bowel preparation only.

IEL and IEM were released from biopsies using DTT/EDTA and harvested by centrifugation at 300 *g* (5 min). IEM were obtained by centrifugation of resulting supernatants at 4500 *g* (20 min). LPL were obtained by digestion with collagenase/liberase of remaining tissue. All cells were washed, then phenotyped and counted by flow cytometry. Cells were first stained for viability using LIVE/DEAD fixable-near-IR stain (ThermoFisher) before addition of surface-staining antibodies in fetal calf serum. Cells were stained with antibodies to γδ TcR, CD4, CD8, CD103, CD39, and CD73 (Supplementary Table 1). All samples were acquired on a BD Biosciences FACS Canto II flow cytometer and data analyzed by FlowJo software (Tree Star).

### Commensal-specific T and B cell memory proliferative responses

CD4/CD8 T cell and B cell proliferative responses were measured in PBMC in response to the following nine species: *Bacillus licheniformis, Bacteroides ovatus, Bifidobacterum pseudocatenulatum, Clostridium paraputrificum, Escherichia coli, Hafnia paralvei, Schaalia turicensis, Staphylococcus epidermidis* and *Veillonella atypica*. The species were selected from an initial panel of 19 on the basis of their relatively high immunogenicity^16^. They were isolated from the cecum of healthy donors ^17,18^ and grown anaerobically in Hungate tubes containing Wilkins-Chalgren broth (37 °C for 24 h). Aliquots (1 ml) were centrifuged at 13,000 rpm for 10 min, supernatants removed and cell pellets snap-frozen with dry ice before storage at -80 °C. PBMC were obtained over Ficoll gradients and labelled with CellTrace Violet™ (1 µM, ThermoFisher) according to manufacturer’s instructions, then cultured at 4×10^6^/ml in XVIVO15 serum-free medium (Lonza, + 50 µg/ml gentamycin (Sigma) and penicillin/streptomycin (ThermoFisher, 1/100)). Killed bacteria (2×10^5^, enumerated by SYBR Green staining and flow cytometry) were added to 0.2 ml PBMC cultures and microbe-specific proliferation in gated populations determined after seven days culture. Cultured cells were analyzed by staining with LIVE/DEAD stain and CD4/CD8/CD19 antibodies (Supplementary Table 1). Positive responses were classed as those showing >2% divided cells and at least twice the unstimulated control level.

### Measurement of antibody-bound microbes

IEM were labelled with SYBR Green DNA stain (ThermoFisher, 1/100,000), anti-IgA-APC/anti-IgG-APC/Cy7 and analyzed by flow cytometry to determine proportions (%) of bacteria coated with antibodies in the gut. Intact microbes were gated according to SYBR Green staining and total proportions of bacteria staining positive for IgA and IgG determined.

### Analysis of circulating B cell subsets

We examined naïve vs memory and effector memory B cell subsets, along with IgA- and IgG-switched B cells, plasmablasts and transitional B cells (T1 and T2), in live PBMC using antibodies listed in Supplementary File 1. We previously published an example of the gating strategy used^4^. CD19^+^ B cells were classified into memory (CD27^+^ IgD^-^), effector memory (CD27^-^ IgD^-^), plasmablast (CD27^hi^ CD38^hi^), T1 transitional (CD24^hi^ CD38^hi^ within the naïve (CD27^-^ IgD^+^) and T2 transitional (CD24^lo^ CD38^lo^ within naïve) subpopulations, expressed as percentages of the total B cell fraction. B cells expressing surface IgA and IgG were also analyzed.

### Statistical analysis

GraphPad Prism 9 software (GraphPad, San Diego, CA) was used to plot and analyze the data, using Mann Whitney non-parametric tests to compare groups. P values less than 0.05 were considered significant and indicated by: *:p<0.05; **:p<0.01; ***:p<0.001; ****:p<0.0001.

## Results

### γδT cells with residence markers and CD8^+^ Trm are deficient in FAP IEL

IEL consisted of γδ T cells and CD8^+^ T cells. In healthy controls the majority of these expressed CD103 and were therefore classed as tissue resident in phenotype or Trm. In FAP samples CD103 expression was greatly reduced in both populations (Figure 1). The reduction in the proportion of CD103-expressing cells (of total live cells) was 94-97% for CD8 T cells and 90-93% for γδ T cells (Figure 1b). This was predominantly due to a loss of CD103 expression rather than a depletion of total T cells (Figure 1c).

**Figure 1.**
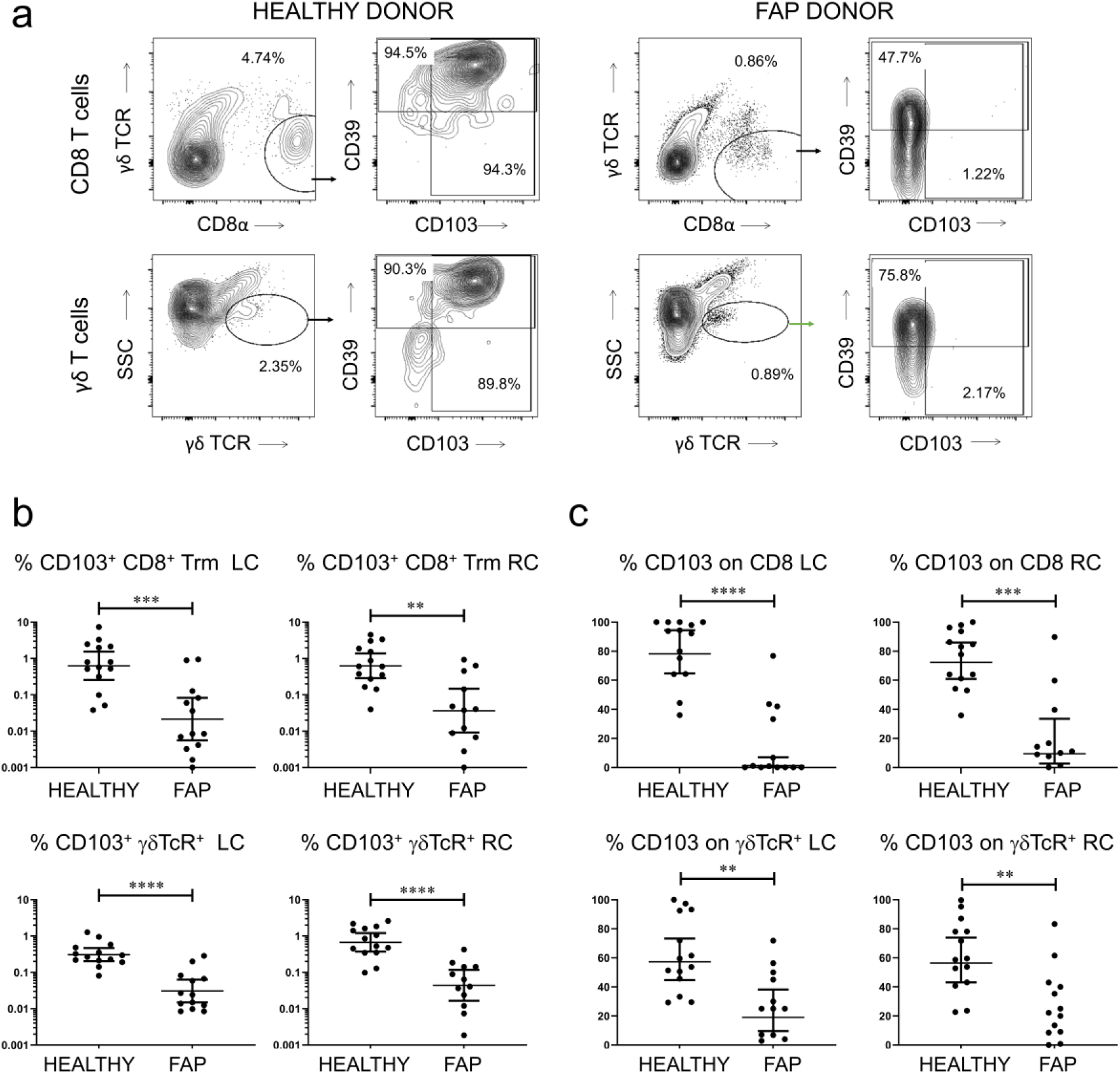
Deficiency of resident memory T cells and γδ T cells in FAP IEL. **a**: Comparison of IEL staining profiles from representative healthy control (left) and FAP (right) colonic biopsies, reflecting loss of CD103 but not CD39 expression on CD8^+^ IEL (upper panels) and γδ T cells (lower panels). Similar results were obtained from left and right colon. **b**: Pooled data from all donors showing percentages of CD8^+^γδ TCR^-^ CD103^+^ Trm as a proportion of total IEL, in left colon (LC) and right colon (RC). **c**: Proportions of cells expressing CD103 within gated CD8 and γδ T cells, as in b. Median values ± 95% confidence intervals are shown; statistically significant differences between groups (Mann Whitney tests) are indicated.

### CD4^+^ and CD8^+^ Trm are deficient in FAP LPL and CD4 Trm lose CD73 expression

We performed a similar analysis of T cells in LPL but examined CD4^+^ and CD8^+^ αβ T cells, since CD4 T cells are largely absent from IEL and γδ T cells rare in LPL. The results indicated further significant reductions in Trm in mucosa, with 83-85% reduction in CD8 Trm and 74-77% fewer CD4 Trm, with significant loss of CD103 expression on both populations in the left colon (Figure 2a). Due to the higher yield of LPL vs IEL, we were able to accurately assess both CD39 and CD73 expression on CD4 and CD8 T cells in LPL. Consistent with our previous study^16^, CD103^+^ T cells preferentially expressed CD39, a regulatory T cell-associated ectonucleotidase not present on circulating T cells. Expression of this marker was not lost on T cells in FAP, although there was a trend towards reduced expression in CD8 T cells. By contrast, CD73 expression was significantly downregulated on CD4 T cells but not CD8 T cells in both left and right colon (Figure 2b). This was unexpected since CD39 but not CD73 is preferentially expressed on CD103-expressing T cells in the colon, with CD73 being generally highly expressed.

**Figure 2.**
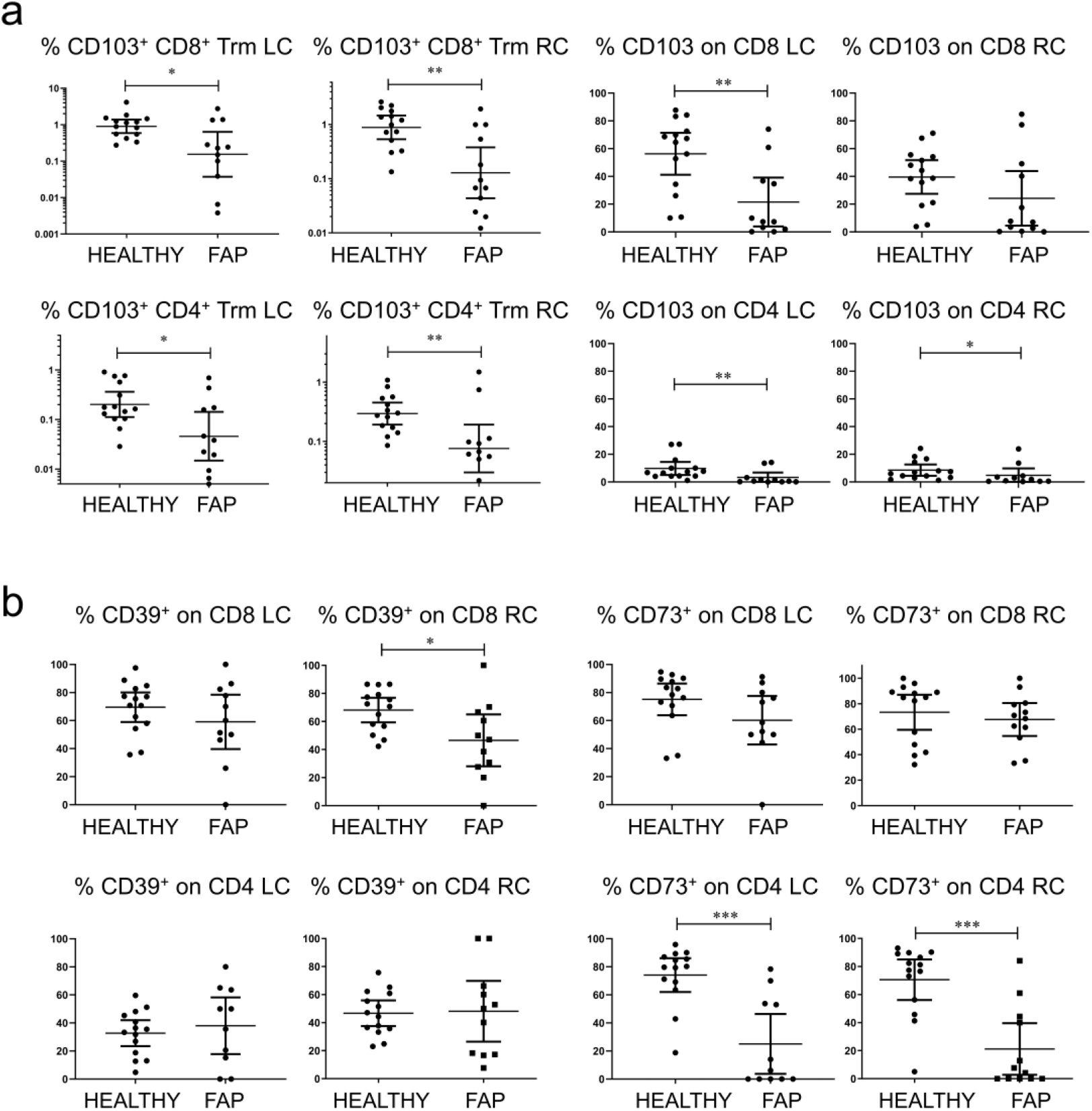
Reduced CD103 and CD73 expression on lamina propria T cells in FAP. **a**: Percentages of CD8^+^ (upper panels) and CD4^+^ (lower panels) CD103^+^ Trm, as proportions of total LPL (left) in left and right colon (LC, RC) of healthy vs FAP donors. Right-hand graphs show proportions expressing CD103 within gated T cell fractions. **b**: Expression of regulatory T cell markers CD39 and CD73 on CD4 and CD8 Trm in LPL. Median values ± 95% confidence intervals are shown; statistically significant differences between groups (Mann Whitney tests) are indicated.

### IgA response to intestinal microbiota is enhanced in FAP

Intraepithelial microbes were released from colonic biopsies and assessed for coating with IgA and IgG. The data revealed significantly higher levels of IgA coating in IEM from right colon of patients with FAP compared to healthy controls (Figure 3), a finding not seen in CRC^4^. A similar trend was apparent in the left colon but did not reach statistical significance. The IgG levels present on these microbes varied greatly from individual to individual in both groups, but there was no significant difference between healthy controls and patients with FAP.

**Figure 3.**
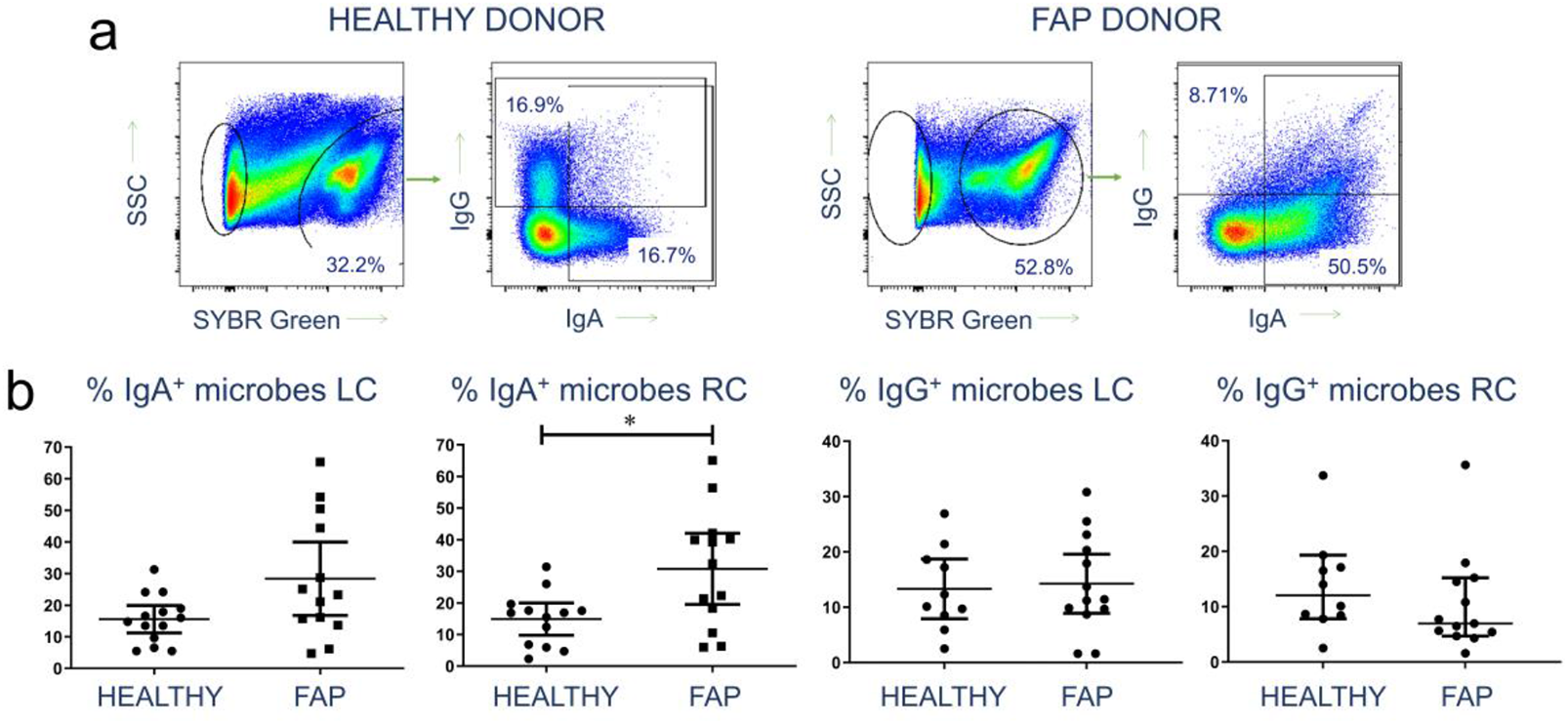
Increased IgA response to intraepithelial colonic microbes in FAP. **a**: Representative flow cytometry profiles showing identification of IEM by DNA staining, followed by analysis of IgA and IgG antibody coating of bacteria, in a healthy control (left) and a FAP donor (right). **b**: Pooled data from all donor left and right colon (LC, RC) samples, showing proportions of IEM coated with IgA and IgG. Median values ± 95% confidence intervals are shown; statistically significant differences between groups (Mann Whitney tests) are indicated.

### Enhanced levels of IgA-switched B cells and T1 transitional B cells in blood of patients with FAP

Since mucosal antibody production to the microbiota appeared raised in FAP, and we had observed signs of B cell activation in blood from CRC patients in a previous study^4^, we performed B cell subset analysis on PBMC from both groups. The data (Fig 4) showed a similar but weaker pattern to CRC, with significantly more T1 transitional B cells than healthy controls, but unlike CRC^4^, no significant change in effector memory B cells or plasmablasts. Consistent with the data on IEM coating, IgA^+^ and not IgG^+^ B cells were increased in blood of FAP patients, in contrast to CRC where neither subset was altered^4^.

**Figure 4.**
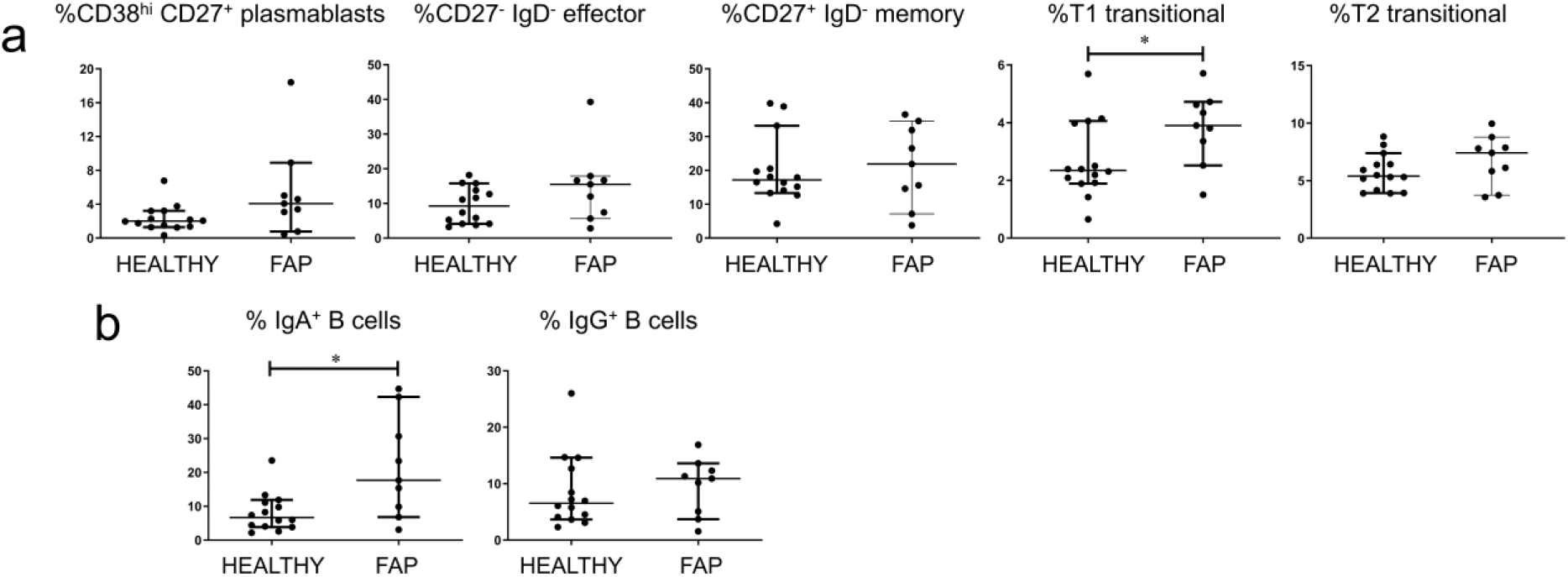
Increase in circulating IgA-switched B cells and T1 transitional cells in FAP. **a**: Representation of different maturational stages of B cells within total circulating CD19^+^ cells in healthy control and FAP blood. Cells were gated as described in Materials and Methods. **b**: Proportions of IgA- and IgG-switched B cells in blood, as a proportion of the total CD19^+^ population, determined as in a. Median values ± 95% confidence intervals are shown; statistically significant differences between groups (Mann Whitney tests) are indicated.

### CD4 T cell memory to commensal bacteria is enhanced in FAP

Since IgA responses to the microbiota were enhanced in FAP compared with controls, we analyzed T and B cell memory to a panel of commensal intestinal bacteria, to determine whether specific species were involved in the disease process. As we previously observed in other studies^16,4^, proliferative responses were observed in CD4, CD8 and CD19 populations in all donors, with highly individual patterns of specificity and strongest memory apparent in CD4 T cells (not shown). The strongest CD4 T cell memory responses in patients with FAP were to *Hafnia paralvei* and *Clostridium paraputrificum*, whilst the weakest CD4 memory was to *Staphylococcus epidermidis* and *E. coli*. The magnitude of the proliferative responses could not be compared between healthy controls and patients with FAP since there was a significant age difference between the two groups (Table 1), and magnitude of response correlated negatively with age. Therefore, we compared the numbers of positive responses (Fig 5) seen to the panel of bacteria, which did not correlate with age. Further, no age correlation was seen in any other data presented in Figs 1-4. Only CD4 T cell responses showed a statistically significant difference, with patients with FAP having more memory responses than healthy controls.

**Figure 5.**
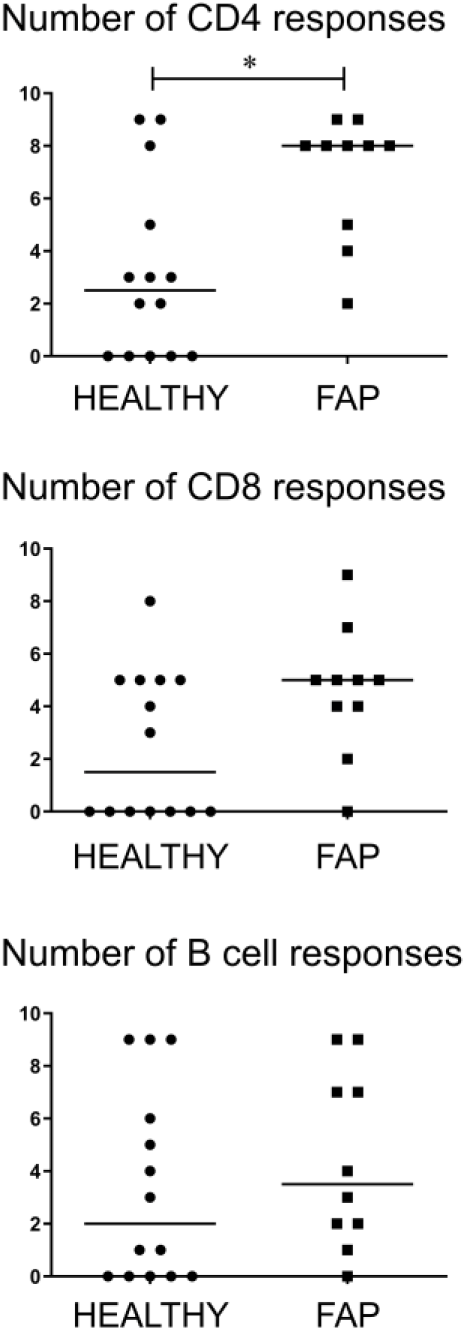
Increased CD4 T cell memory to commensal bacteria in FAP. Proliferative responses of gated CD4^+^, CD8^+^ and CD19^+^ T/B cells in PBMC were determined in response to a panel of 9 commensal bacterial species (listed in Materials and Methods). Numbers of positive responses within the panel were defined as those with >2% cell division and >2 times background proliferation. Median values ± 95% confidence intervals are shown; statistically significant differences between groups (Mann Whitney tests) are indicated. The magnitude of proliferation to each species is not included as the data were biased by donor age difference between groups.

## Discussion

Patients with FAP showed a marked deficiency in CD8 and CD4 resident memory T cells in colonic tissue - populations that promote barrier immunity and have regulatory functions. The deficiency appeared to be mainly due to loss of CD103 expression rather than depletion of total T cells in the tissue. CD103 is a defining marker of Trm since it mediates tethering of T cells to epithelium^19^, promoting long-term residence in the mucosa and tumor tissue^20^. The findings are consistent with a study of *APC* mutant mice, which found lowered proportions of CD8^+^ T cells in small intestine lamina propria and lower CD8 T cell IFN-γ production from Peyer’s patches^21^. Interestingly, FAP tissue has been shown to have altered retinoic acid metabolism, with reduced RALDH1A1/2 expression in adenomatous epithelia relative to healthy tissue and increased CYP26A1 in diseased crypts of patients with FAP^22^. Retinoic acid is critical for induction of CD103 and gut homing of T cells^23^.

Since Trm represent a large fraction of the immune system and maintain barrier immunity against the intestinal microbiota, the loss of such cells is likely to contribute to the inflammatory events that precede carcinogenesis. Furthermore, patients with FAP showed a marked deficiency in γδ T cells in epithelial tissue - cells with a unique ability to kill tumor cells^24, 25^. Increased IgA, but not IgG responses to epithelial microbes were observed, suggesting increased bacterial translocation into colonic tissue driving mucosal antibody secretion. This could be a direct result of weakened immune barrier function. Overall, our data support the hypothesis that poor retention of tissue residence and altered immunity to commensal bacteria may contribute to dysbiosis and result in poor immune surveillance for tumor cells, exacerbating the extremely high CRC risk in FAP. This is consistent with the lack of observed ileal adenomas or ileal cancer in FAP, given that this tissue has a lower bacterial load than the colon and higher lymphoid infiltration. It is possible that medications may have altered the microbiota in some patients. However, pre-surgical antibiotics were administered within the 24 hours prior to surgery so may not have impacted mucosal populations.

Catabolism of extracellular ATP is a key pathway in immune regulation which controls the activation of dendritic cells, the inducers of acquired immunity in the intestine following sampling of luminal contents or infection^26, 27^. CD39 and CD73 are ectonucleotidases key to ATP catabolism to ADP and adenosine, respectively. Both markers are highly expressed on Foxp3-expressing regulatory T cells and also on human Trm^16, 28^. ATP released by microbes and activated T cells^29, 30^ needs to be broken down by ectonucleotidases or it will activate dendritic cells^31^. The loss of CD73 expression on CD4 Trm in FAP colonic tissue is a novel finding, but is consistent with animal model data implicating the *APC* gene in T cell function^32^. Murine studies often focus on the role of intestinal Foxp3^+^ CD4 regulatory T cells^10^, but these represent a smaller proportion of human lamina propria T cells^16^. Whether CD4 Trm from human FAP colonic mucosa have altered regulatory T cell function, as a result of reduced CD73 expression, should be a topic for further research.

When we examined peripheral blood, lymphocytes from patients with FAP demonstrated a CD4 T cell activation and IgA signature, while CRC shows a strong B cell activation signature without IgA^4^. It is difficult to speculate on the reasons for these differences in immune profile, since the CRCs we previously studied were not the result of FAP. However, development of the majority of colorectal adenomas and carcinomas does involve somatic mutations of *APC*^33^. The increased T1 transitional B cells in FAP may reflect altered B cell selection, since they are immature B cells with enhanced autospecificity. Although FAP is rare compared to CRC, our current data throw light on immune dysregulation pathways that could lead to CRC. Interestingly, the increased CD4 response to commensals in our FAP cohort was not selectively directed to *E. coli*, a species reported to invade the mucosa in FAP^15^. Instead, the responses showed highly varied specificities and magnitudes in each patient, as observed in healthy control donors. FAP patients had CD4 T cell memory to more commensal species than healthy controls, suggesting that poor barrier function was allowing increased presentation of commensal antigens in mucosal-associated lymphoid tissue, which could directly lead to increased IgA secretion into the lumen. It should be noted, however, that healthy donors display readily detectable CD4 memory to many commensals, and that the apparent immune tolerance to intestinal microbiota is therefore localized to the mucosa. The globally altered pattern of immunity in patients with FAP might be easily exploited by the more invasive species identified in the study of Dejea et al^15^, which may be able to colonize the mucosa without inducing an abnormally strong response.

Overall, our study has important clinical implications in FAP and points towards future novel strategies for managing the disease. For example, stimulation of intestinal retinoic acid production might recruit Trm, promoting immune surveillance and preventing tumor cell invasiveness. Understanding possible alterations in ATP catabolism within FAP tissue could also yield valuable research directions. Immunotherapies that promote intestinal T cell regulatory functions and IL-10 production could be used to control inflammatory pathways leading to CRC. Finally, altering the microbiota through dietary intervention or manipulating anti-commensal immunity through vaccination might be capable of correcting immune dysregulation in FAP and other disease states.

## Data Availability

Data can be requested from the corresponding author

## Acknowledgements

We thank Alison Scoggins for administrative assistance. We are grateful to the staff and patients of St Mark’s Hospital and our blood donors for their participation in this study.

## Abbreviations

FAP: familial adenomatous polyposis
APC: adenomatous polyposis coli
CRC: colorectal cancer
Trm: resident memory T-cells
IBD: inflammatory bowel disease
IEL: intraepithelial lymphocytes
LPL: lamina propria lymphocytes
IEM: intraepithelial microbes.

## References

1. Wang T, Cai G, Qiu Y, et al. Structural segregation of gut microbiota between colorectal cancer patients and healthy volunteers. ISME J. 2012;6(2):320–9. doi:10.1038/ismej.2011.109.

2. Tjalsma H, Boleij A, Marchesi JR, Dutilh BE. A bacterial driver-passenger model for colorectal cancer: beyond the usual suspects. Nat Rev Microbiol. 2012;10(8):575–82. doi:10.1038/nrmicro2819.

3. Crucitti A, Corbi M, Tomaiuolo PM, et al. Laparoscopic surgery for colorectal cancer is not associated with an increase in the circulating levels of several inflammation-related factors. Cancer Biol Ther. 2015;16(5):671–7. doi:10.1080/15384047.2015.1026476.

4. Noble A, Pring ET, Durant L, et al. Altered immunity to microbiota, B cell activation and depleted γδ / resident memory T cells in colorectal cancer. MedRxiv. 2021; https://doi.org/10.1101/2021.02.15.21251750.

5. Miyoshi Y, Ando H, Nagase H, et al. Germ-line mutations of the APC gene in 53 familial adenomatous polyposis patients. Proc Natl Acad Sci U S A. 1992;89(10):4452–6. doi:10.1073/pnas.89.10.4452.

6. Kinzler KW, Vogelstein B. Lessons from hereditary colorectal cancer. Cell. 1996;87(2):159–70. doi:10.1016/s0092-8674(00)81333-1.

7. Nieuwenhuis MH, Vasen HF. Correlations between mutation site in APC and phenotype of familial adenomatous polyposis (FAP): a review of the literature. Crit Rev Oncol Hematol. 2007;61(2):153–61. doi:10.1016/j.critrevonc.2006.07.004.

8. van de Wetering M, de Lau W, Clevers H. WNT signaling and lymphocyte development. Cell. 2002;109 Suppl:S13–9. doi:10.1016/s0092-8674(02)00709-2.

9. Smits R, van der Houven van Oordt W, Luz A, et al. Apc1638N: a mouse model for familial adenomatous polyposis-associated desmoid tumors and cutaneous cysts. Gastroenterology. 1998;114(2):275–83. doi:10.1016/s0016-5085(98)70478-0.

10. Aguera-Gonzalez S, Burton OT, Vazquez-Chavez E, et al. Adenomatous Polyposis Coli defines Treg differentiation and anti-inflammatory function through microtubule-mediated NFAT localization. Cell Rep. 2017;21(1):181–194. doi:10.1016/j.celrep.2017.09.020.

11. Akeus P, Langenes V, von Mentzer A, et al. Altered chemokine production and accumulation of regulatory T cells in intestinal adenomas of APC(Min/+) mice. Cancer Immunol Immunother. 2014;63(8):807–19. doi:10.1007/s00262-014-1555-6.

12. Zhou L, Sonnenberg GF. Essential immunologic orchestrators of intestinal homeostasis. Sci Immunol. 2018;3(20)doi:10.1126/sciimmunol.aao1605.

13. Amsen D, van Gisbergen K, Hombrink P, van Lier RAW. Tissue-resident memory T cells at the center of immunity to solid tumors. Nat Immunol. 2018;19(6):538–546. doi:10.1038/s41590-018-0114-2.

14. Nizard M, Roussel H, Diniz MO, et al. Induction of resident memory T cells enhances the efficacy of cancer vaccine. Nat Commun. 2017;8:15221. doi:10.1038/ncomms15221.

15. Dejea CM, Fathi P, Craig JM, et al. Patients with familial adenomatous polyposis harbor colonic biofilms containing tumorigenic bacteria. Science. 2018;359(6375):592–597. doi:10.1126/science.aah3648.

16. Noble A, Durant L, Hoyles L, et al. Deficient resident memory T cell and CD8 T cell response to commensals in inflammatory bowel disease. J Crohns Colitis. 2020;14(4):525–537. doi:10.1093/ecco-jcc/jjz175.

17. Hoyles L, Murphy J, Neve H, et al. Klebsiella pneumoniae subsp. pneumoniae-bacteriophage combination from the caecal effluent of a healthy woman. PeerJ. 2015;3:e1061. doi:10.7717/peerj.1061.

18. Hoyles L, Jimenez-Pranteda ML, Chilloux J, et al. Metabolic retroconversion of trimethylamine N-oxide and the gut microbiota. Microbiome. 2018;6(1):73. doi:10.1186/s40168-018-0461-0.

19. Schon MP, Arya A, Murphy EA, et al. Mucosal T lymphocyte numbers are selectively reduced in integrin alpha E (CD103)-deficient mice. J Immunol. Jun 1 1999;162(11):6641–9.

20. Gauthier L, Corgnac S, Boutet M, et al. Paxillin binding to the cytoplasmic domain of CD103 promotes cell adhesion and effector functions for CD8+ resident memory T cells in Ttumors. Cancer Res. 2017;77(24):7072–7082. doi:10.1158/0008-5472.CAN-17-1487.

21. Tanner SM, Daft JG, Hill SA, Martin CA, Lorenz RG. Altered T-cell balance in lymphoid organs of a mouse model of colorectal cancer. J Histochem Cytochem. 2016;64(12):753–767. doi:10.1369/0022155416672418.

22. Penny HL, Prestwood TR, Bhattacharya N, et al. Restoring retinoic acid attenuates intestinal inflammation and tumorigenesis in APCMin/+ mice. Cancer Immunol Res. 2016;4(11):917–926. doi:10.1158/2326-6066.CIR-15-0038.

23. Jaensson-Gyllenback E, Kotarsky K, Zapata F, et al. Bile retinoids imprint intestinal CD103+ dendritic cells with the ability to generate gut-tropic T cells. Mucosal Immunol. 2011;4(4):438–47. doi:10.1038/mi.2010.91.

24. Ma R, Yuan D, Guo Y, Yan R, Li K. Immune Effects of gamma delta T cells in colorectal cancer: A Review. Front Immunol. 2020;11:1600. doi:10.3389/fimmu.2020.01600.

25. Hayday AC. gamma delta T cell update: adaptate orchestrators of immune surveillance. J Immunol. 2019;203(2):311–320. doi:10.4049/jimmunol.1800934.

26. Schnurr M, Then F, Galambos P, et al. Extracellular ATP and TNF-alpha synergize in the activation and maturation of human dendritic cells. J Immunol. Oct 15 2000;165(8):4704–9.

27. Idzko M, Hammad H, van Nimwegen M, et al. Extracellular ATP triggers and maintains asthmatic airway inflammation by activating dendritic cells. Nat Med. 2007;13(8):913–9. doi:nm1617[pii]10.1038/nm1617.

28. Deaglio S, Dwyer KM, Gao W, et al. Adenosine generation catalyzed by CD39 and CD73 expressed on regulatory T cells mediates immune suppression. J Exp Med. 2007;204(6):1257–65. doi:jem.20062512[pii]10.1084/jem.20062512.

29. Iwase T, Shinji H, Tajima A, et al. Isolation and identification of ATP-secreting bacteria from mice and humans. J Clin Microbiol. 2010;48(5):1949–51. doi:10.1128/JCM.01941-09.

30. Schenk U, Westendorf AM, Radaelli E, et al. Purinergic control of T cell activation by ATP released through pannexin-1 hemichannels. Sci Signal. 2008;1(39):ra6. doi:1/39/ra6[pii]10.1126/scisignal.1160583.

31. Silva-Vilches C, Ring S, Mahnke K. ATP and its metabolite adenosine as regulators of dendritic cell activity. Front Immunol. 2018;9:2581. doi:10.3389/fimmu.2018.02581.

32. Gounari F, Chang R, Cowan J, et al. Loss of adenomatous polyposis coli gene function disrupts thymic development. Nat Immunol. 2005;6(8):800–9. doi:10.1038/ni1228.

33. Powell SM, Zilz N, Beazer-Barclay Y, et al. APC mutations occur early during colorectal tumorigenesis. Nature. 1992;359(6392):235–7. doi:10.1038/359235a0.

