## Supplemental Table 1 for "Dysbiosis in pre-cancerous mucosa may drive immune dysfunction – a study of immune system: microbiota interaction in familial adenomatous polyposis"

Supplementary Table 1. Antibodies used:

| ANTIGEN | FLUOROCHROME | mAb CLONE | SUPPLIER |
|---|---|---|---|
| CD4 | BB515 | SK3 | BD Biosciences |
| CD8α | BV605 | SK1 | BioLegend |
| CD19 | PE-Cy5 | HIB19 | BD Biosciences |
| CD24 | APC-Cy7 | ML5 | BioLegend |
| CD27 | PE | M-T271 | BD Biosciences |
| CD38 | BV605 | HIT2 | BioLegend |
| CD39 | PE-Cy7 | A1 | BioLegend |
| CD69 | BV605 | FN50 | BioLegend |
| CD73 | APC | AD2 | BioLegend |
| CD103 (integrin αE) | PE | Ber-ACT8 | BioLegend |
| IgA | APC | IS11-8E10 | Miltenyi Biotec |
| IgD | PE-Cy7 | IA6-2 | BD Biosciences |
| IgG | FITC | IS11-3B2.2.3 | Miltenyi Biotec |
| TCR γδ | FITC | 11F2 | BD Biosciences |
